# Unlocking Efficient Hospital Operations: Discrete Event Simulation in R

**DOI:** 10.1101/2023.09.21.23295327

**Authors:** Mohamed Kamal, Omneya Hassanain

## Abstract

In healthcare systems, optimizing resource allocation without compromising patient care is paramount. This study employs a simulation-based approach to evaluate the efficiency of bed allocation within a hospital setting. Utilizing a patient arrival model with an exponential distribution, we simulated patient trajectories to examine system bottlenecks, particularly focusing on waiting times. Initial simulations painted a scenario of an “unstable” system, where waiting times and queue lengths surged due to the limited number of available beds. Through iterative simulations, we explored the operational research question: “What is the minimum number of beds required to stabilize the system?” Our results, visualized in a series of detailed metrics plots, suggest that the addition of a specific number of beds can significantly reduce patient waiting time and stabilize the system. This research offers insights for hospital management on resource optimization, potentially leading to improved patient care and reduced operational costs.

## Introduction

Healthcare facilities operating in today’s demanding environment often struggle with several challenges, including a surge in patient numbers, the financial burden of providing quality care, and stringent budget constraints.^1,2^ To assist healthcare decision-makers in optimizing their available resources, modeling and simulation techniques have been employed to assess hospital operations’ efficiency.^3–3^ These approaches not only evaluate the current strategies but also offer insights into the potential outcomes of future modifications to the healthcare system. Models serve as mathematical frameworks that aim to represent specific facets of reality in sufficient detail, enabling the estimation of the consequences of healthcare decisions.^7,8^ They simplify intricate systems by distilling them to their fundamental components, making them indispensable tools in health technology assessment.^9^ Simulation, as a pivotal aspect of this process, involves the computer-based execution of a model. It allows for the in-depth study of system properties, operational characteristics, and the evaluation of various strategies. Decision-makers can draw conclusions and make informed choices based on simulation results before implementing any changes in the actual healthcare system.^10^ Simulation serves as a valuable instrument for enhancing the utilization of existing resources and aids in the development of new strategies by leveraging data collection and information technology.^11^ Simulation of a system refers to the computational emulation of a model, enabling an in-depth examination of its features, operational characteristics, and responses to different strategies.^9^ The process allows for data-driven decision-making by mimicking real-world systems in a controlled environment, thus enabling insights into the efficacy of potential operational strategies prior to their actual implementation. Simulation serves as a critical tool for optimizing resource allocation and planning novel strategies, backed by empirical data and advanced information technology.^10^ The process of modeling can generally be segregated into two main phases:^12^ the first phase is the Conceptualization of the Problem, which involves the transformation of domain-specific knowledge, such as healthcare operations, into a simplified yet representative problem set. It essentially lays the groundwork for simulation by delineating the scope, limitations, and key variables of the problem at hand. The second phase is the Conceptualization of the Model. This phase aims to identify the most appropriate simulation methodology that aligns with the specific requirements and complexities of the problem under scrutiny. This includes the selection of techniques, algorithms, and validation procedures to build a model that can simulate the real-world system effectively. Moreover, several pivotal factors critically impact the model’s architecture and utility. The nature of the problem under investigation dictates the selection of decision alternatives, criteria, and constraints. The target population informs the model’s scalability and the granularity of data required. The time horizon over which outcomes are measured affects the discounting rates and can introduce dynamic complexities. Finally, the project’s overarching objectives inform the key performance indicators (KPIs) and the decision rules embedded in the model. These interrelated components collectively influence not just the model’s design but also the data required for its population, the analytic methods employed, and the format and comprehensibility of model reporting.^13,14^ In healthcare decision analytic models, domain experts wield significant influence over both the initial conceptualization of the problem and the resulting model specifications. The initial problem framing should aim for comprehensive coverage of the healthcare issue at hand, irrespective of the constraints imposed by data availability. Recognizing the ramifications of data quality, or the lack thereof, is crucial. The structure of the model must be scrutinized for its sensitivity to missing or poor-quality data, commonly conducted through sensitivity analyses, to ensure robustness and validity in decision-making.^15,16^ In the realm of healthcare decision analysis, a multitude of modeling techniques exist to facilitate evidence-based decision-making. Among the foundational approaches are decision trees, which serve as graphical representations enumerating possible patient trajectories, each with a quantifiable outcome and associated probability. While decision trees offer a robust framework for scenarios with a limited outcome set and relatively short temporal dimensions, they exhibit limitations when applied to chronic conditions like cancer. In such cases, parameters can be non-stationary over elongated time horizons, time-to-event is a significant variable, and events may recur. Markov models emerge as a superior alternative in these contexts, offering the flexibility to account for long-term projections, variations in event timing, and recurring events.^12,17^ Markov models are cohort-based models on which a hypothetical homogenous cohort of patients moves through defined health states. Time is segmented into cycles, during which patients may either remain in their current health state, transition to a different state, or progress to an absorbing state, typically death, based on specified transition probabilities. As the simulation unfolds, various outcomes—such as Quality-Adjusted Life Years (QALYs) or associated costs accumulate, facilitating comparative evaluation of different intervention strategies. Despite their relative simplicity, cohort-based Markov models are not without limitations. One key constraint is the inherent ‘Markovian assumption’ that transition probabilities are memoryless and independent of past states. While the granularity of health states can be increased to mitigate this issue, this often leads to an exponential growth in model complexity. When the required number of states becomes unwieldy, or when capturing individual-level variability is crucial, individual-based simulation models emerge as a more robust alternative, allowing for a more nuanced representation of patient heterogeneity.^15,18,19^ Discrete Event Simulation (DES) emerges as a potent tool for conducting individual-level simulations. Originating from methodologies in engineering and operations research, DES allows for a sophisticated representation of complex systems. Unlike traditional cohort-based models, DES facilitates the modeling of individual entities as they transition through a series of distinct states. Events, defined by specific triggers or conditions, catalyze these transitions and consequently impact various outcomes. By capturing the idiosyncratic nature of these events and transitions, DES affords a more nuanced and comprehensive understanding of healthcare scenarios.^20,21^ One prominent approach in simulation is Discrete Event Simulation (DES), which replicates the operations of real or proposed systems as a series of ordered events. Each event signifies a specific change in the system’s state at distinct time points. DES empowers decision-makers to construct intricate operation models, swiftly assess “what-if” scenarios, and explore alternative methods for implementing new strategies.^22^ DES It excels in replicating the dynamic behaviors of intricate systems and the interactions among individuals, populations, and their surroundings.^23^ The primary objective of such modeling is to comprehensively evaluate various practices or strategy options, particularly in situations where conducting necessary surveys or comparative experiments in real-world settings may be impractical. In contrast to aggregate models that lack interaction, such as decision trees or Markov models,^10^ DES, as an operational research technique, offers distinct advantages by modeling complex systems at the individual level rather than the cohort level. The scenarios simulated are often conceptualized as queuing networks, where individual entities progress through a sequence of discrete events at discrete intervals. During the intervals, they may wait in queues due to limited resource availability.^24^ DES also provides decision-makers with the ability to perform “what if” analyses by modifying operational scenarios and rules. This enables them to anticipate potential impacts resulting from various policy alternatives before implementing changes in the existing systems. DES is often the simulation modality of choice under certain conditions. Specifically, DES is preferable when the model necessitates a vast array of states, involves competitive allocation of resources leading to queue formations, or requires the modeling of complex interactions between individual entities. However, it’s important to acknowledge that DES models, like other modeling techniques, present simplified representations of reality. Central to DES are six core concepts: entities, attributes, events, resources, queues, and time. Entities—often represented as patients in healthcare scenarios—possess attributes that modify the likelihood of specific event occurrences. These entities vie for limited resources, resulting in the formation of queues which can be managed through various service disciplines, such as ‘First-In, First-Out’ or priority-based arrangements. Attributes of these entities, such as age, gender, health status, medical history, disease duration, and other demographics, are updatable while the models are running.^25,26^ Events in DES encompass a wide range of occurrences during simulation, including the onset or recurrence of diseases, admission to healthcare facilities, delivery of medical treatments, or transitions between health states. Events in a simulation model can encompass a diverse array of phenomena relevant to healthcare outcomes. These may include the onset or recurrence of a medical condition, admission to a healthcare facility, administration of therapeutic interventions, or transitions between distinct health states. These events serve as key markers or milestones within the simulation, facilitating a more nuanced understanding of healthcare dynamics. The inherent flexibility of DES allows these events to occur, and recur, in any logical sequence, thereby capturing the complexities of real-world healthcare systems.^25,26^ One of the principal advantages of DES is its discrete time-handling mechanism. The simulation advances in time solely at ‘discrete’ junctures where events transpire, eliminating the need for intermediate calculations. This results in significantly expedited computational processing, making DES a computationally efficient simulation technique. The objective of this manuscript is to elucidate a methodological framework aimed at facilitating informed decision-making among healthcare decision-makers. This methodology involves the modeling of patient trajectories, resource allocation, and other pertinent parameters within healthcare facilities. Additionally, it entails simulating various scenarios and generating user-friendly outputs, particularly tailored for executive management. Within the scope of this paper, we undertake an illustrative example of the implementation of modeling and simulation techniques, which serve the overarching purpose of mitigating wait times and reducing queues within a healthcare environment. The hypothetical scenario posits a situation characterized by a finite number of available beds set against a surging rate of inpatient admissions, thereby resulting in the emergence of extensive waiting lists. To address this predicament, we propose employing Discrete Event Simulation (DES) as an analytical tool to devise effective solutions.

### Stages of DES Modeling Process

The stages of DES modeling process are outlined in figure (1). The first step in developing DES is to define the system under investigation and relevant events that can occur to individuals in that system. This is assisted by performing a model structure in which all possible scenarios and their required parameters are identified. The next step is to estimate those parameters using pre-collected data, if required data are not available or of a poor quality the expert in the system operation may be consulted for data elicitation and their responses should be validated. If expert elicited data are not of appropriate validity, the model should serve only as a guide for further data collection needed to build the model. Model implementation involves transferring conceptual model structure into a computer program. The model can be populated and analyzed using DES dedicated software which may offer a better visualization of the problem through animations and also ease of application by a non-expert. Another approach is the use of a general programming language such as “R”. The selection between general-purpose programming languages and specialized DES software presents a trade-off. General-purpose languages, such as ‘R’, offer greater flexibility in system representation, reduced simulation run times, and independence from commercial software licenses. However, these advantages come at the cost of a steeper learning curve, necessitating the development of bespoke code for implementing package functions like event lists, queue formation, resource utilization, and probabilistic sampling. This, in turn, demands rigorous code debugging. Model output includes both mean values and distributions of values, which align with the specific objectives of the study. To assess the stability of this output, it is commonly recommended that the variation between different model runs should be less than 5%. In some specific settings, this threshold may be adjusted to as low as 1%. To enhance output stability, several approaches can be utilized, such as increasing the number of entities participating in the simulation, extending the model’s temporal horizon, or increasing the number of replications within a single model run. The method selected for improving stability will depend on the unique characteristics of the system under study.^27–27^ In order to ascertain the fidelity of a simulation model to the empirical reality it seeks to emulate, rigorous validation techniques must be rigorously applied. Such techniques are instrumental in confirming the model’s capacity to furnish accurate and reliable outputs that can be generalized to the system under investigation. Concurrently, sensitivity analysis serves as an indispensable tool for model optimization. It allows for the comprehensive assessment of the impact of varying input parameters on model outcomes, thereby identifying variables of significant influence. Through this bifurcated approach of validation and sensitivity analysis, the model not only gains credibility but also reaches an optimized state that enhances the robustness of its predictive and prescriptive capabilities.^30–30^ For disseminating Discrete Event Simulation (DES) research findings, both general and detailed perspectives should be presented to provide a comprehensive overview of the model’s architecture and underlying logic as such the reporting on DES studies should seamlessly blend general overviews with detailed insights to enable a thorough understanding of both the model’s structure and its methodological intricacies.^32^ The general perspective offers a high-level view that encapsulates the main elements of the model, such as states, events, and flow mechanisms. This facilitates an initial, holistic understanding of the model’s purpose and functionality. Conversely, the detailed perspective delves into the minutiae of specific states, transition probabilities, and event sequences. This level of granularity is critical for experts who may wish to replicate the model, execute sensitivity analyses, or validate the model against empirical data. Supplementing these perspectives, it is crucial to include event documentation figures that itemize the sequence of events, their associated attributes, and their implications for model outcomes. Such documentation serves multiple functions, from explicating the model’s internal dynamics to facilitating its interpretation, replication, and external validation by experts in the field.

**Figure (1):**
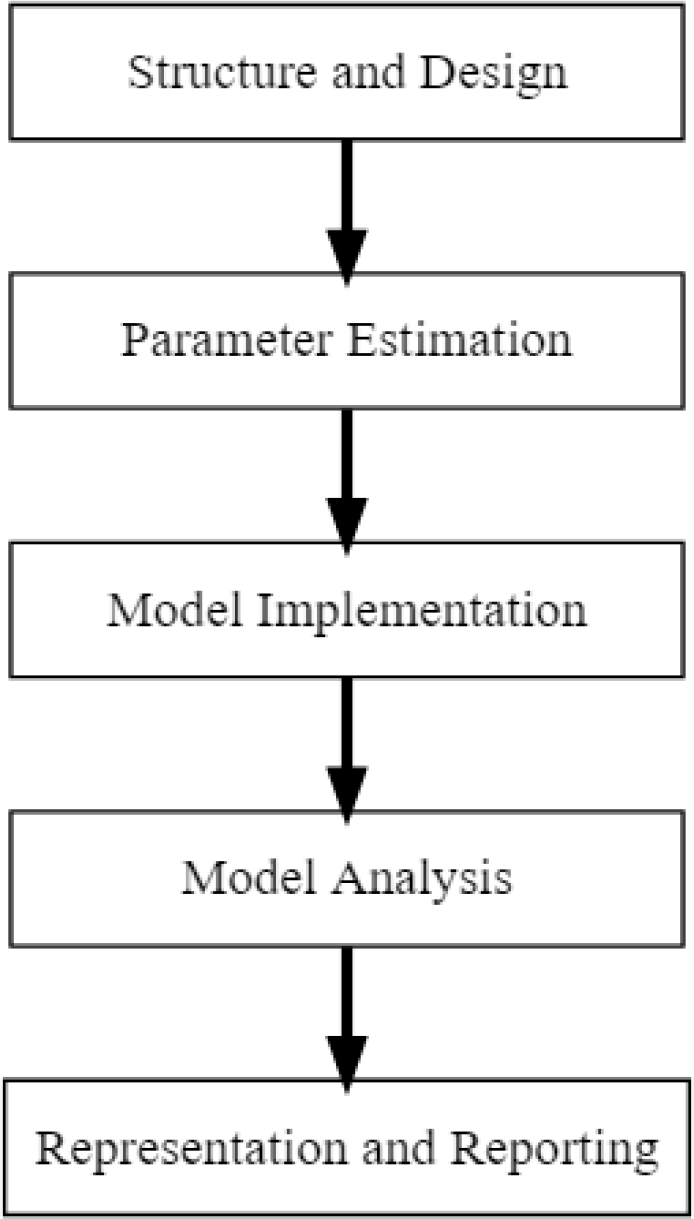
Stages of DES modeling process

## Methodology

In this article we used R version 4.2.1 and simmer_4.4.6.2.^33,34^ For our hypothetical example, we assume that we have collected relevant data and estimated data distributions for all variables needed to construct the model, for example, Inter-arrival time, Length of Stay (LOS) for each phase, etc. For model validation purposes, the user needs to also collect outcome variables like distribution of waiting time, waiting queue, etc. Assume you have a hospital with 30 inpatient beds allocated to accommodate patients of a specific disease. Let us assume that all patients accommodated will be treated by a standard protocol composed of 4 treatment phases. Before writing the code, we started by drawing the decisions tree containing the probabilities, inter-arrival time (IAT), distributions and priorities of entities to compete for resources (beds in our case). A simplified example is shown in figure (2). Practically, it can get much more complicated with nested branches and other add-on attributes. The next step is to model the workflow along with relevant parameters and attributes into the system.

**Figure (2).**
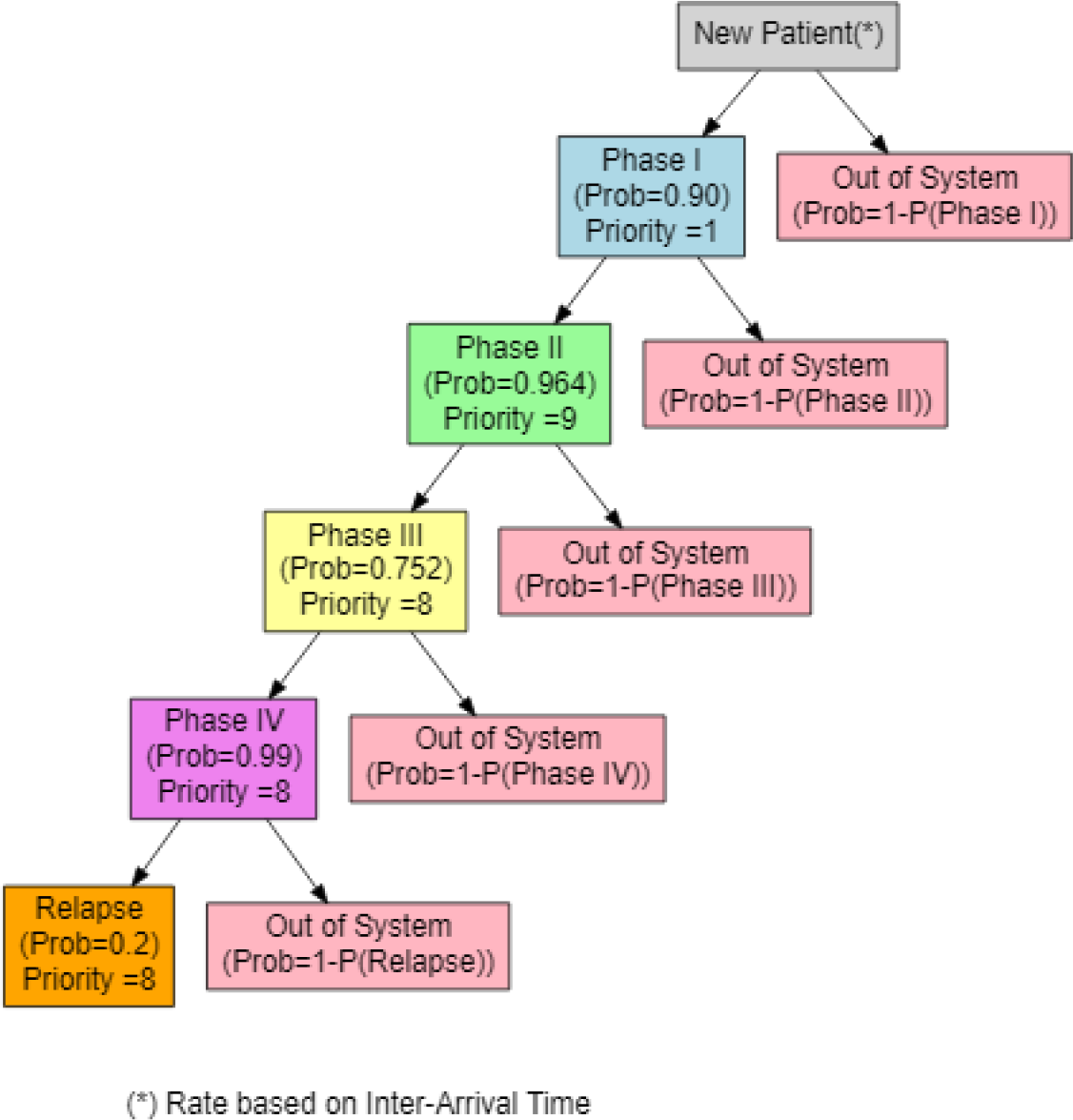
Decision tree for the treatment protocol milestones with probabilities and priorities.

**Figure (3).**
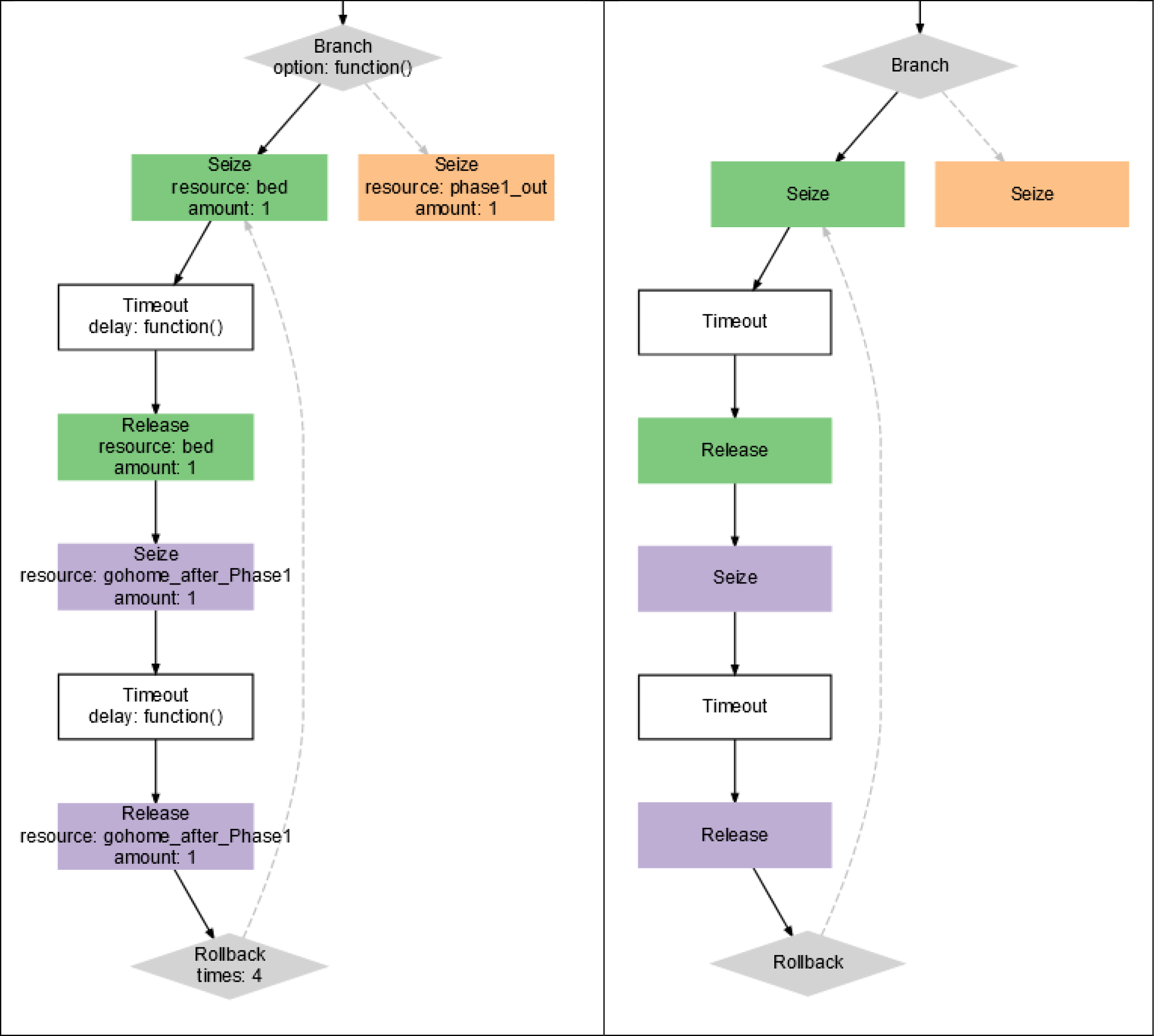
Comparison of patient trajectory plots generated by simmer: on the left with the ‘verbose=TRUE’ setting and on the right with ‘verbose=FALSE’. This showcases the level of detail controlled by the verbose argument.

**Figure (4).**
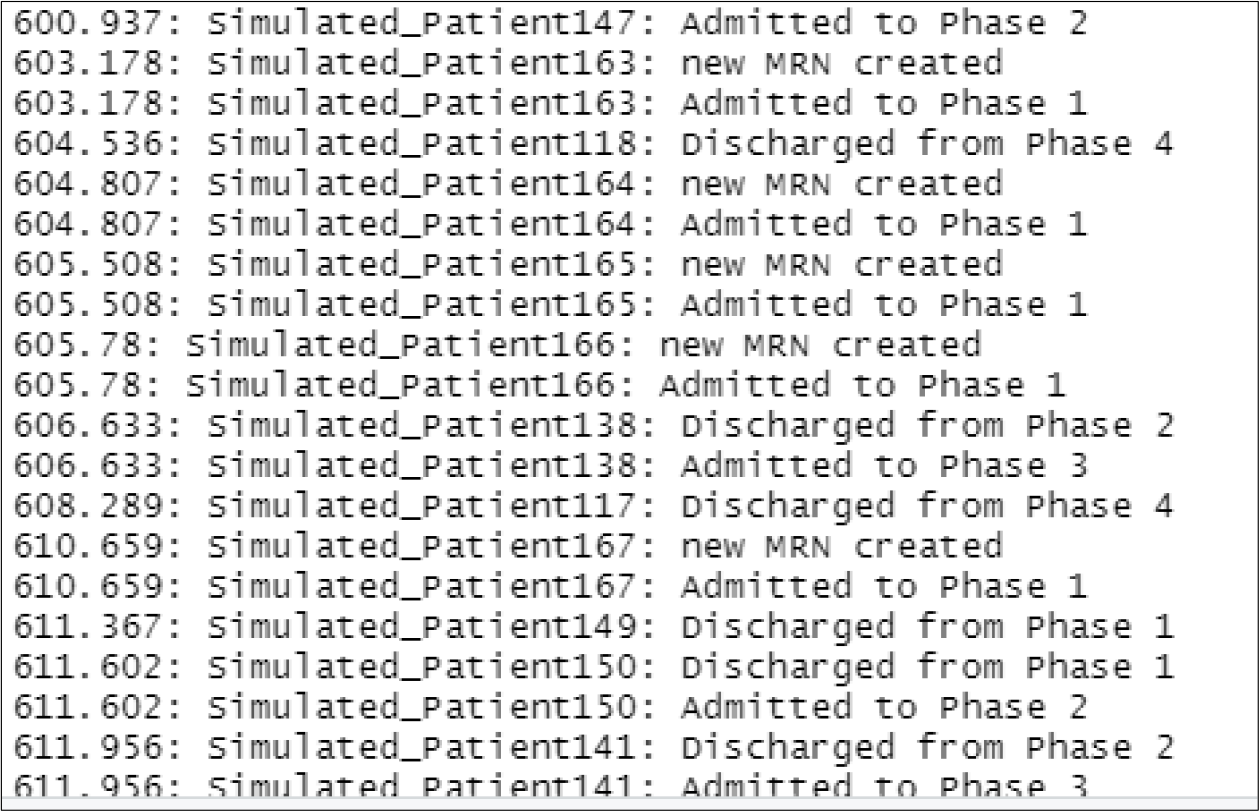
DES in progress, displaying simulation time, simulated patient identifier, and the milestones.

The R code snippet, R Code (1), is an example for modeling patient trajectories through different phases of treatment for a disease (referred to as “DiseaseX”). The code uses the simmer package to set up a discrete-event simulation environment. Parameters Setup sets up the number of beds (nbeds), repetitions (myrep), simulation period (period), and Inter Arrival Time (myIAT) as initial parameters for the simulation. A simulation environment (env) is initialized where all the actions take place. A trajectory for each patient, named “DiseaseX_patient_path”, is created which consists of multiple treatment phases (Phase1, Phase2, etc.) and possible relapses. To demonstrate how to add attributes to entities, each patient gets a health attribute using rnorm(), but in a real scenarios replace it with a relevant equation or other distributions. The patients go through different phases like Treatment_Phase1, Treatment_Phase2, etc. At each phase, the availability of a bed is checked, waiting time calculated followed by using the resource, (seize(“bed”, amount = 1)), and a treatment timeout from the pre-specified distribution is implemented. After each phase, the patient may be discharged or move to the next phase based on probabilistic decisions (branch()). Resources like beds were created using (add_resource(“bed”, nbeds)) and dummy resources for counting purposes were added to track the patient flow. Generators are set up to manage the arrivals of patients using an estimated distribution. The code utilizes add_generator to introduce new patients into the system based on the specified inter-arrival times. While logs and other monitored attributes are captured and saved for further analysis, the script saves various monitored attributes and resources to .csv files. The script also extracts relevant statistics from the logs using text mining. Moreover, specific log patterns can be identified using grep() to extract the number of relapses, new patients, and queues for each repetition to gain extra mining insights. The code computes and saves statistics like Average Length of Stay (LOS) and waiting times as well as detailed logs that are generated for each patient’s journey, capturing entry and exit for each phase. The simulation is executed ‘myrep’ times to achieve a set of simulations results. These multiple runs account for uncertainties in the model, parameters, or input data. Since individual runs might yield results influenced by specific conditions or assumptions, using a multiple runs approach provides a more reliable understanding of the modeled system by examining the range of outcomes. The patient trajectory can be plotted in a more visual friendly way. Below is a simplified patient trajectory output by the system, however, in real-life trajectories can be much more complicated. You can control the level of details in the output plot using the verbose = TRUE / FALSE argument in plot() function.

### Model Testing and Validation

A crucial step in model validation involves aligning model parameters with real-world values to produce outputs that closely resemble actual situations. For this purpose, adjust the model to reflect the current number of beds, patient inter-arrival times, length of stay (LOS), waiting times, and other relevant metrics. Execute the model with an optimal range of simulation repetitions, typically between 1,000 to 10,000 .^35^ This optimal range is chosen because it is sufficiently large to accommodate uncertainties, yet it ensures that computational resources and simulation time are used efficiently without excess.

### Running the Simulation

To highlight the capabilities of our simulator, we deliberately tweaked the parameters to emulate an extended waiting list for patients. Once we’ve validated that the model reflects a hypothetical scenario consistent with the actual status quo, we observe the emergence of an “unstable” system. This instability leads to an accumulation of entities. Figure 5 provides a comprehensive visualization of this instability, capturing: Resource usage over time, Resource utilization, Waiting time, Attributes, and Length of Stay (LOS).

**Figure (5).**
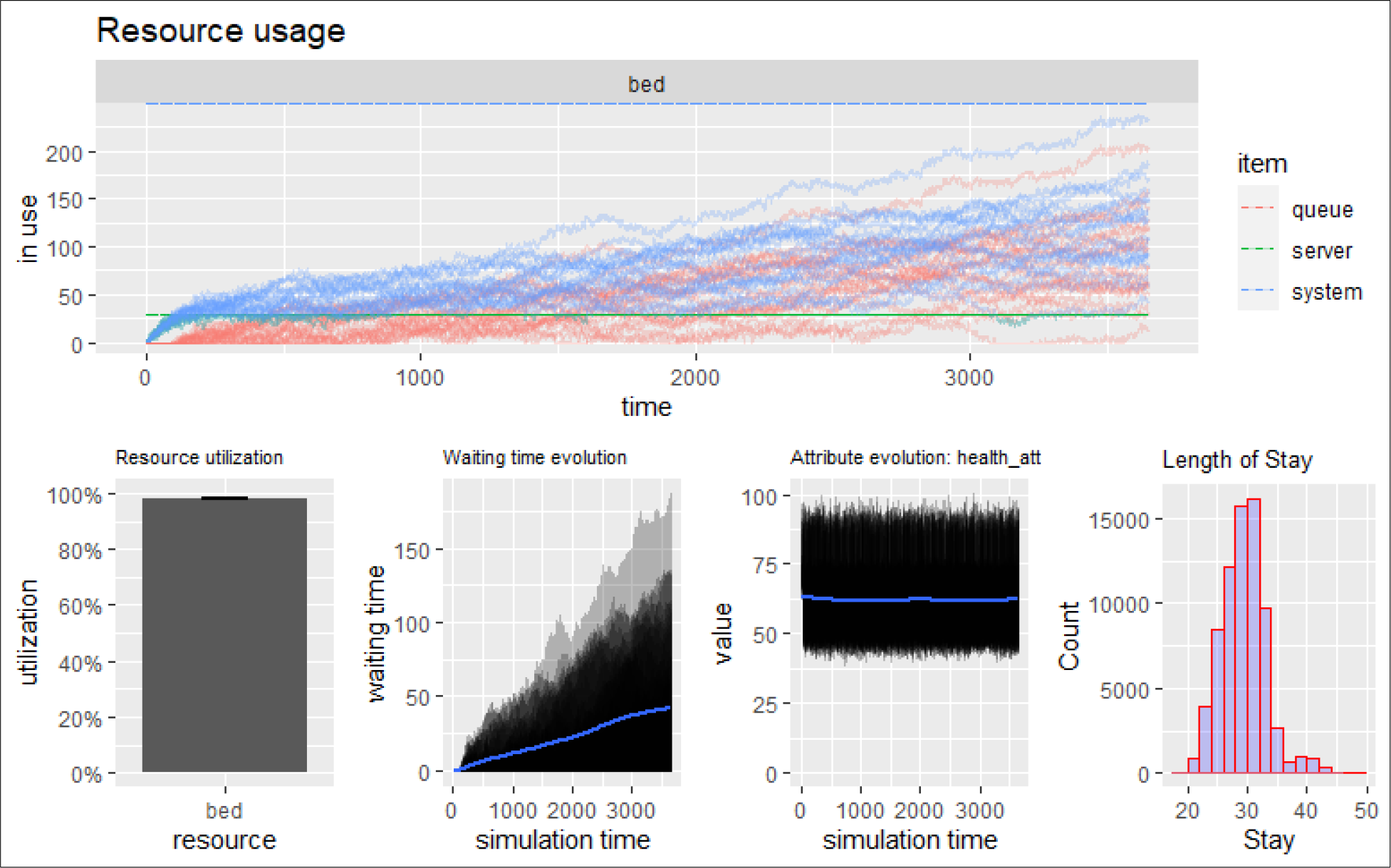
Simulation with 30 Beds: For simplicity, only 20 replications are depicted. The system’s instability leads to an escalating queue and extended waiting times, resulting in a noticeable buildup of patient waiting queues.

Having presented this report to executive management, the pressing challenge arises: how many additional beds are necessary to optimize patient service? While adding more beds incurs substantial costs, the pivotal operational research question becomes: what is the least number of beds required to rectify the situation? To make an informed decision, we ran simulations incrementing bed count by 1, 2, 3, 4, and 5 from our initial resource. Figure 6 offers a closer look at the outcomes of these simulations. Figure 7 provides a comprehensive visualization of the stabilized system after adding 5 extra beds, capturing: Resource usage over time, Resource utilization, Waiting time, Attributes, and Length of Stay (LOS).

**Figure 6.A.**
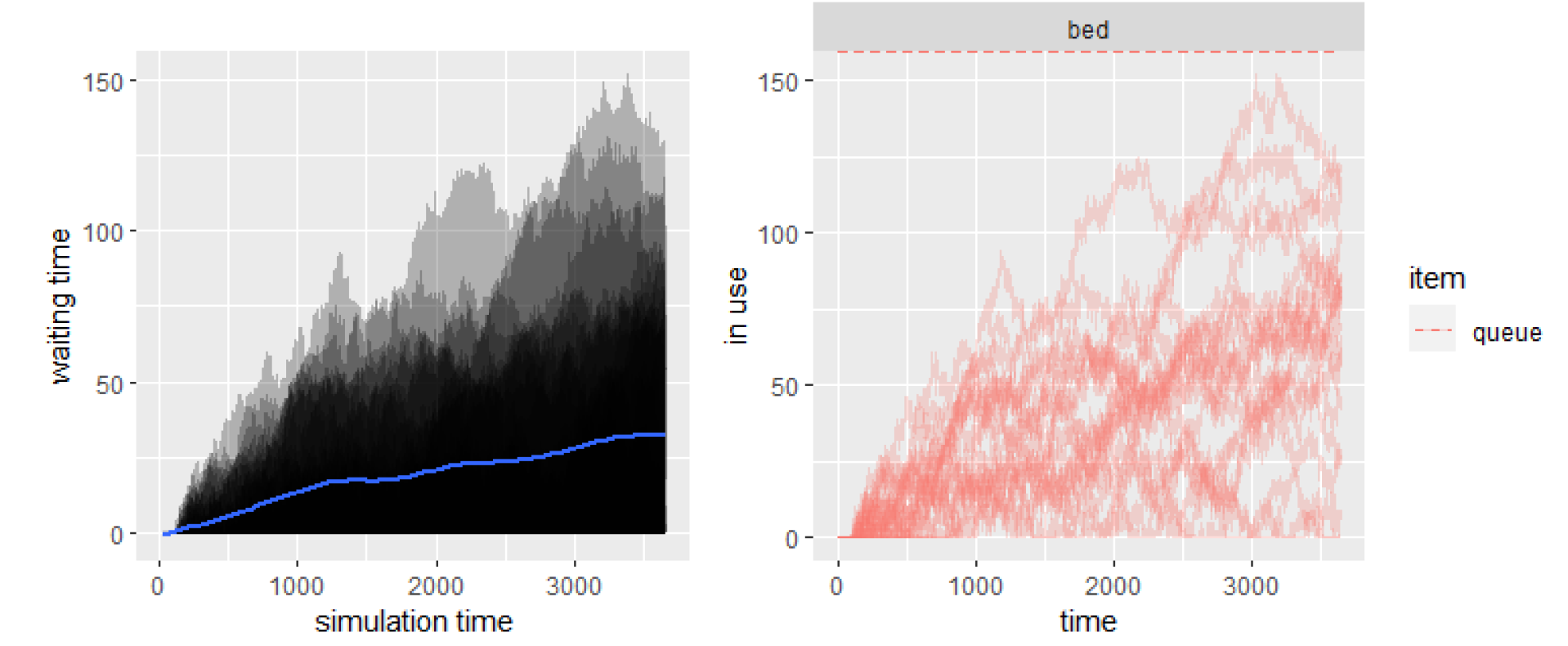
Simulation with 30 beds over 3,650 days using 20 replications. The average queue length is 35, with a mean waiting time of 33 days.

**Figure 6.B.**
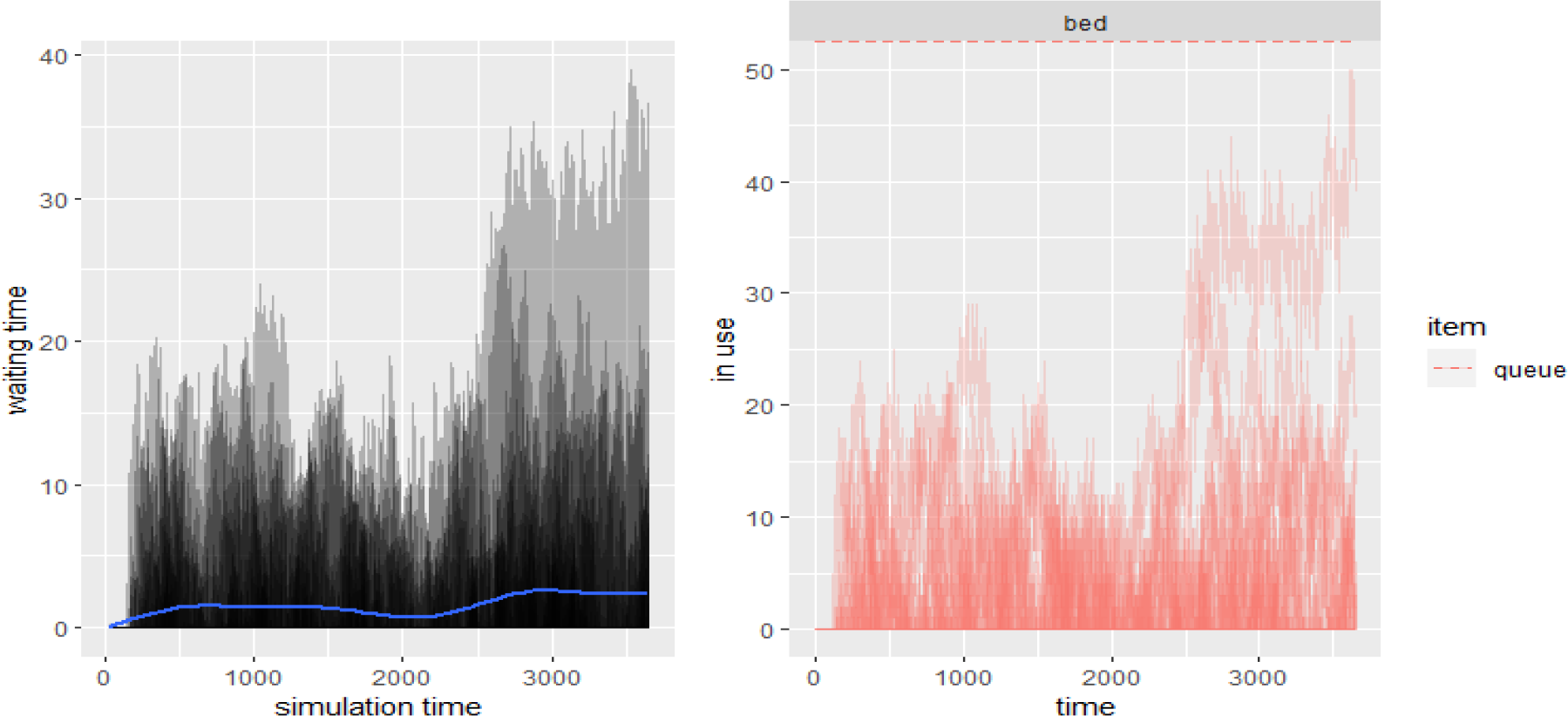
Simulation with 35 beds showing a notable improvement with both the queue length and waiting time averaging 3.

**Figure (7).**
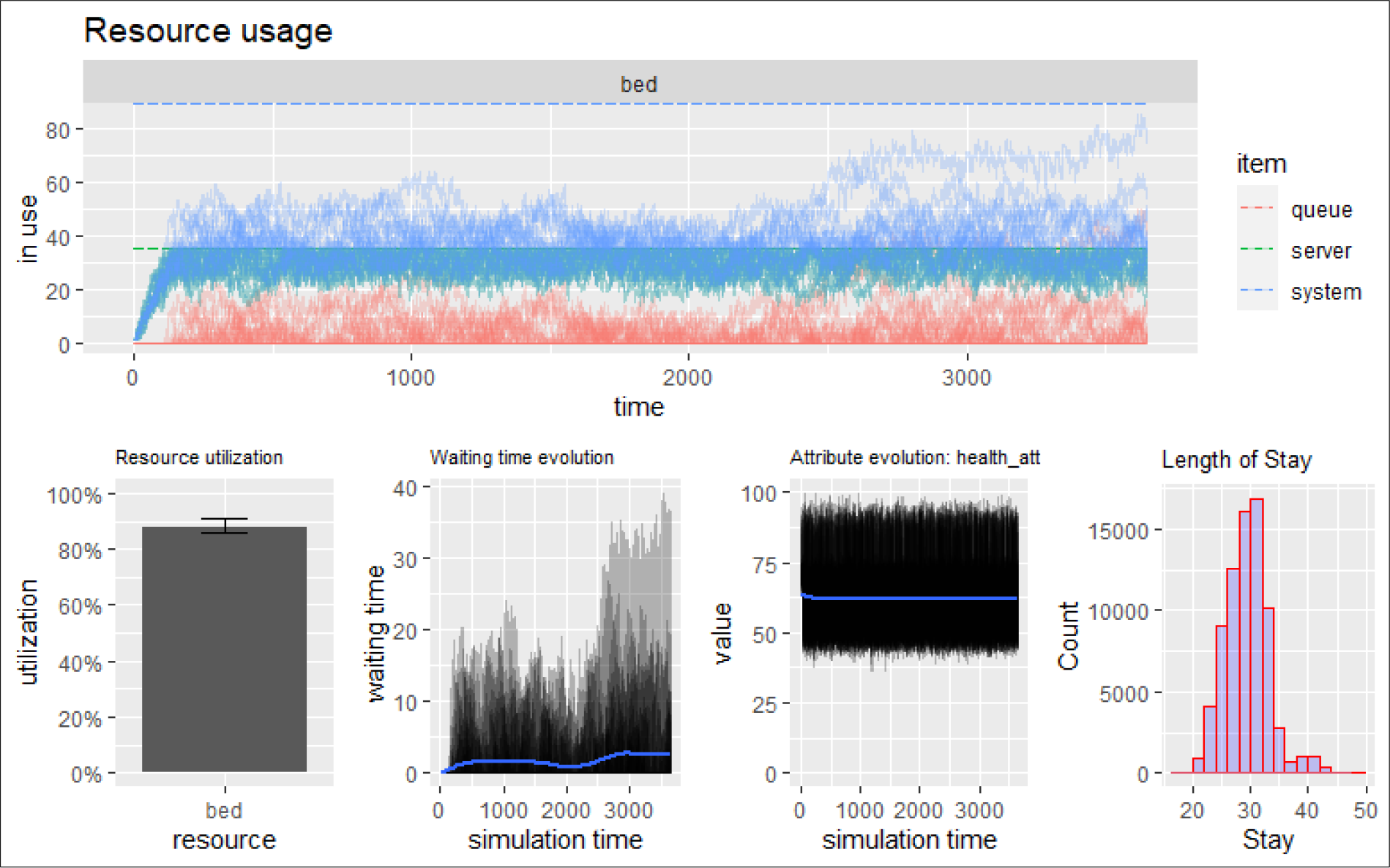
Comprehensive visualization of the system stabilized with an addition of 5 extra beds, resulting in a total of 35 beds. The metrics presented include Resource Usage over Time, Resource Utilization, Waiting Time, Attributes, and Length of Stay (LOS). Notably, queue length and waiting time both average to 3.

## Conclusion

Discrete Event Simulation (DES) stands as an invaluable analytical tool in healthcare decision-making, particularly in understanding, predicting, and optimizing complex systems like patient queues in healthcare facilities. The fundamental advantage of employing DES lies in its flexibility, allowing decision-makers to model, adjust, and re-run scenarios, effectively enabling them to anticipate the outcomes of various system changes before their actual implementation. Through the hypothetical scenario examined in this article, we highlighted the profound impacts of resource constraints, in this case, limited bed availability, on the effectiveness and efficiency of healthcare delivery. By using R and the simmer package, we constructed a comprehensive DES model that not only offers insights into the current state of a system but also provides potential solutions to challenges such as rapidly increasing patient queues. The illustrative example, which simulated the addition of beds in a constrained facility, elucidates how small changes can greatly affect patient waiting times and overall system stability. This ability to run “what-if” scenarios assists executive management in making informed decisions that are both cost-effective and patient-centric. Furthermore, our detailed explanation of the DES modeling process serves as a roadmap for researchers and practitioners looking to implement such simulations in their own contexts. Emphasis on thorough model validation ensures that these models are both credible and adaptable to the multifaceted and ever-evolving dynamics of real-world healthcare environments. In closing, as healthcare systems worldwide struggle with challenges of limited resources and growing demand, tools like DES provide a beacon of hope. By allowing stakeholders to simulate and foresee potential outcomes, DES assists in crafting solutions that are not just reactive but proactive, paving the way for a more efficient and equitable healthcare future.

## Data Availability

This study employs modeling and simulation techniques based on a fictitious example; it does not utilize real-world data. The data is generated through simulation.

### Appendix

#### I. R code (1): Modeling Workflow and Simulation Parameters Input

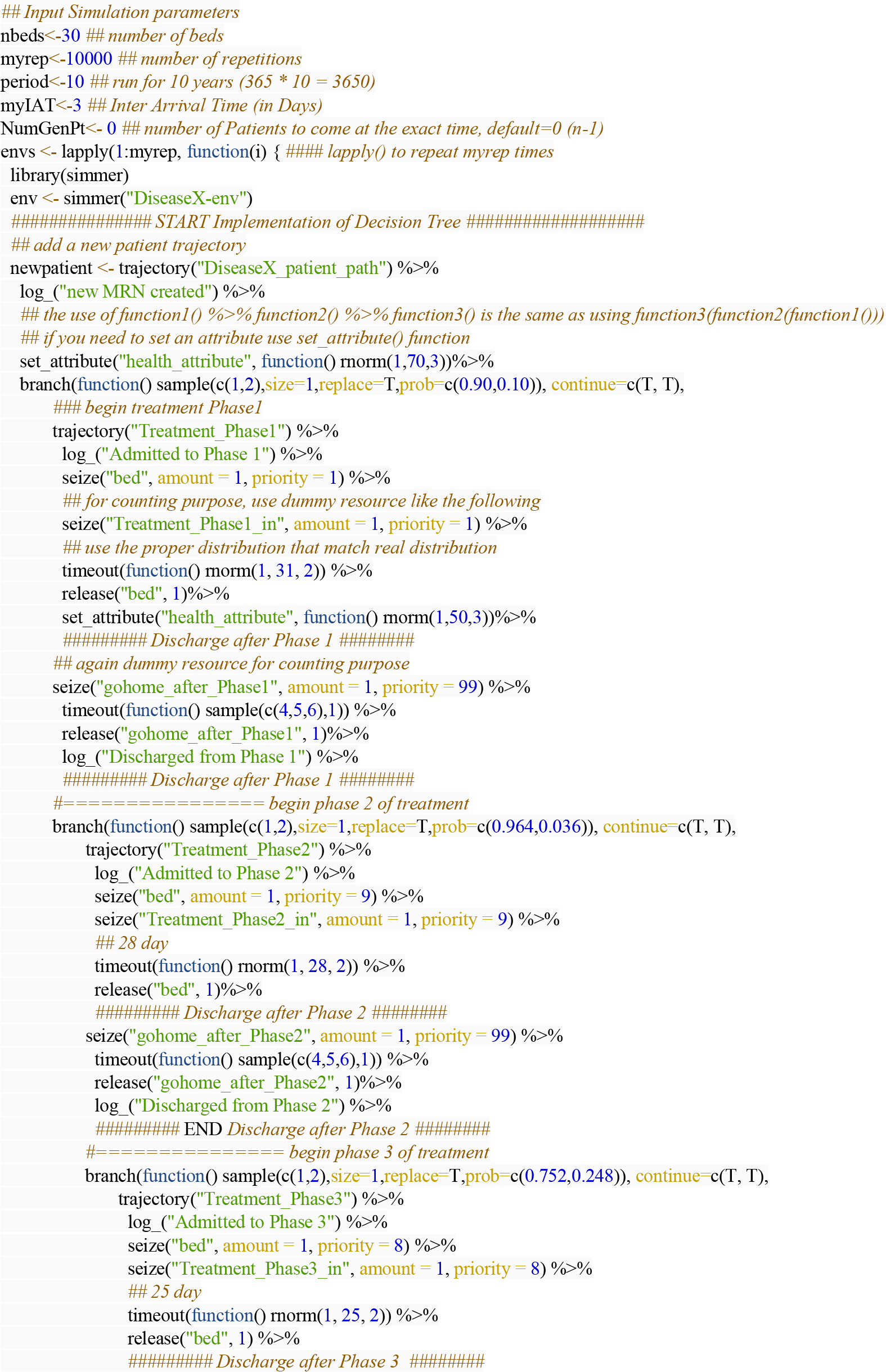

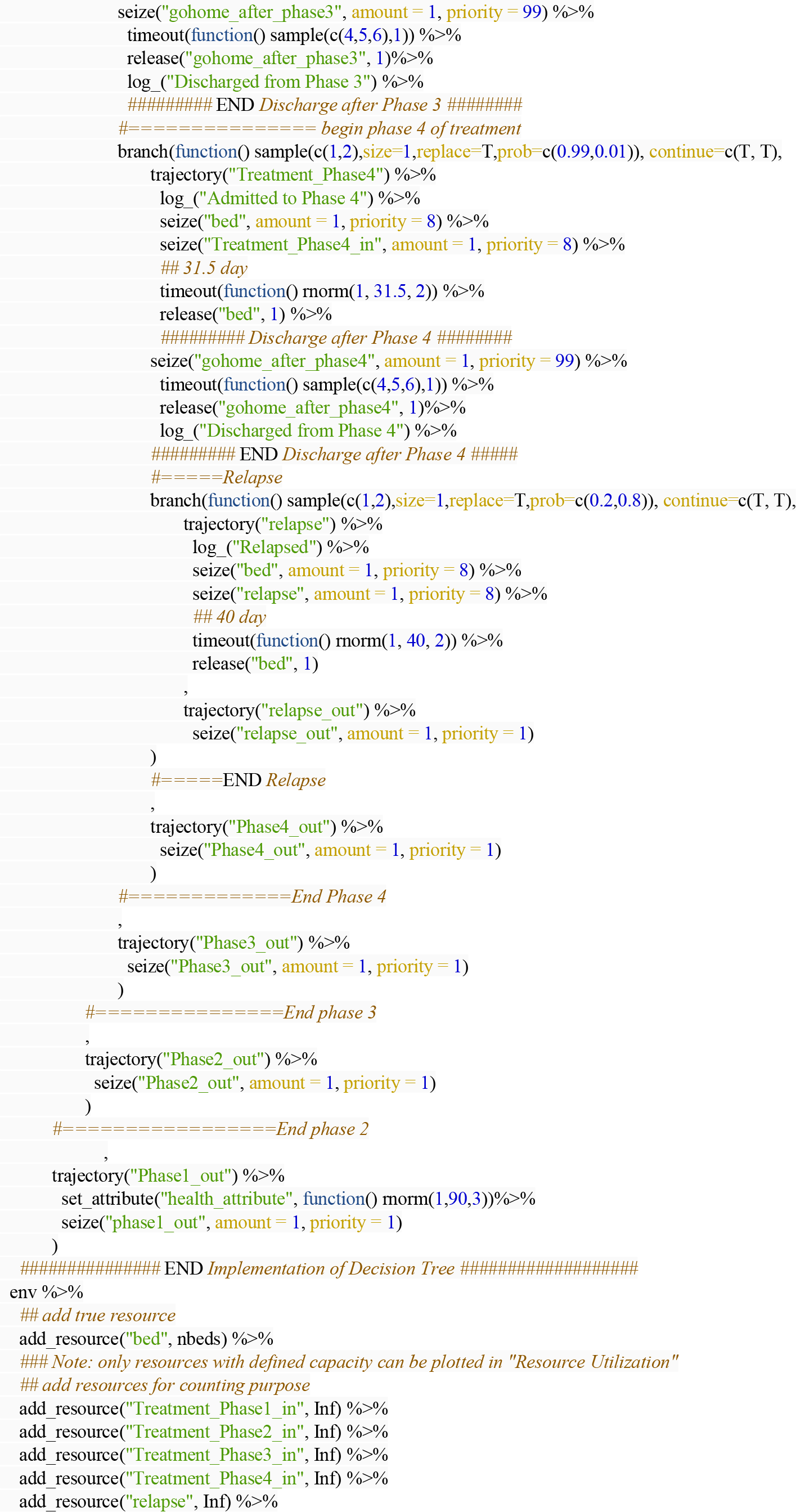

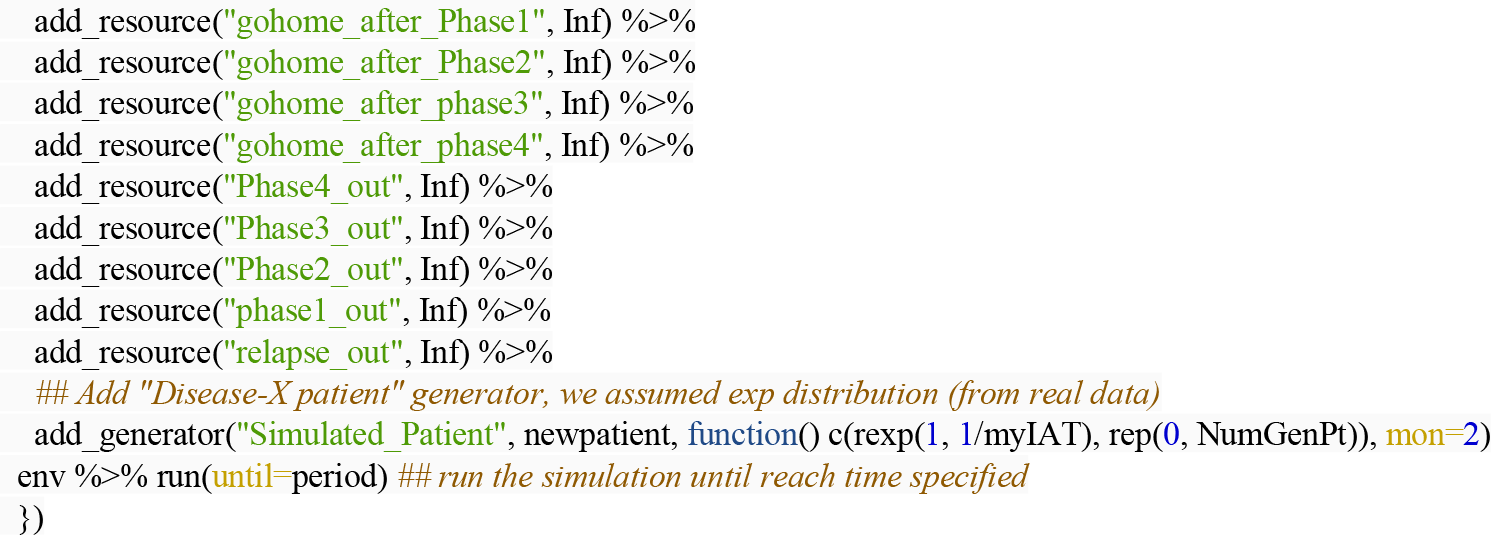

#### II. Session Information

**> sessionInfo()**

R version 4.2.1 (2022-06-23)

Running under: Ubuntu 20.04.4 LTS

attached base packages:

[1] stats graphics grDevices utils datasets methods base

other attached packages:

[1] simmer_4.4.6.2

loaded via a namespace (and not attached):

[1] compiler_4.2.1 magrittr_2.0.3 tools_4.2.1 Rcpp_1.0.8.3

[5] codetools_0.2-18

